# Assessing feasibility and effectiveness of point-of-care limited HPV genotype tests in cervical cancer screening: a modelling study

**DOI:** 10.1101/2025.03.13.25323880

**Authors:** Kyra H. Grantz, Sarah Girdwood, Angela Muriuki, Sergio Carmona, Brooke E. Nichols, Shaukat Khan

## Abstract

**Background:** Cervical cancer (CaCx) remains a significant cause of morbidity and mortality, especially among women in sub-Saharan Africa, where only 15% of women have undergone screening. To address these substantial screening gaps, the WHO recommends human papillomavirus (HPV) DNA detection as the primary screening method. One strategy to improve screening coverage is the use of limited HPV genotype target tests which can be offered as true point-of-care (POC) tests. However, understanding the conditions under which limited-genotype tests can compete with existing full genotype testing remains a barrier to their development and adoption. We explored the trade-offs between accessibility, retention, costs, and test performance to inform the development of new POC limited-genotype tests.

**Methods:** We developed a once-off HPV screening model to investigate the potential use-cases and required specifications of limited-target POC tests. We estimated the proportion of cases of cervical intraepithelial neoplasia grade 2+ (CIN2+) and the testing costs per case identified for three scenarios: (1) limited-genotype tests alone, (2) full-genotype tests, and (3) limited-genotype rule-in tests followed by full-genotype testing for those screening negative. We compared screening program performance across ranges of screening coverage, genotype target capture, loss to follow-up, and test price to identify the conditions where limited-genotype screening was non-inferior to full-genotype screening.

**Results:** When coverage and retention following full-genotype screening were high, limited-genotype tests were only competitive at a very low cost. However, with either modest increases (≥5%) in screening coverage or higher loss to follow-up after full-genotype screening (≥19.8%), an 8-target limited-genotype test could identify the same number of CIN2+ cases as full-genotype screening, but must cost <US$8.50 to be cost-equivalent. Adding full-genotype testing following a rule-in 4-target test would be as effective as full-genotype screening in the number of CIN2+ cases identified; the limited-genotype test must cost <US$2.20 to be cost-equivalent.

**Conclusions:** Our modeling supports screening with an 8-target limited-genotype HPV POC test priced at <US$8.50 for cost-equivalence with full-genotype screening. A rule-in 4-target test will be cost-equivalent if priced at <US$2.20. These insights can enhance CaCx screening access and support the goal of eliminating CaCx in sub-Saharan Africa.

## Background

Cervical cancer remains a significant cause of morbidity and mortality, particularly among women in Africa, where there are an estimated 117,000 new cases and 77,000 deaths due to cervical cancer each year(1). Cervical cancer is a preventable disease and the existence of high performing vaccines against human papillomavirus (HPV) and technologies for screening and treating precancerous cervical lesions make elimination possible. Yet, substantial screening gaps pose a challenge to achieving elimination targets, as only 15% of women in sub-Saharan Africa have ever undergone cervical cancer screening(2). The World Health Organization (WHO) has recently recommended the use of HPV DNA detection as a primary screening method for cervical cancer, replacing visual inspection with acetic acid (VIA) and cytology(3). To ensure they are on trajectory to elimination, it is recommended that countries need to screen 70% of eligible women with a high-performance test at least once by the age of 35 by 2030, and twice in a lifetime(4).

Currently, the WHO recommendation for HPV DNA screening is for a test that includes up to 14 high-risk HPV genotypes (“full-genotype” test). However, this comes at a significant cost compared to the current standard of care, which in many countries is VIA (assay costs of US $10 for centralized full-genotype test or US $15 for near point-of-care (POC) test versus around a US $1 for VIA consumables) contributing to the slow uptake and adoption of HPV molecular screening technologies in Africa(3,5). Further, even when available, the reliance on centralized laboratories for HPV molecular testing can result in limited access and poor retention throughout the diagnostic and care cascade. Shifting testing closer to patients through POC testing technologies could offer a more accessible and feasible alternative to centralized testing. POC HPV testing would enable screen-positive and high-risk women to receive their results in real time, allowing them to be linked to treatment in a single clinical visit, which is particularly valuable in settings with limited healthcare infrastructure. The placement of POC devices in facilities lacking sample referral or with long turnaround times can improve access(6) and retention.

One strategy to improve cervical cancer screening coverage and retention, therefore, is the use of limited HPV genotype target (”limited-genotype”) tests which can be offered as true POC tests with results available within the same clinical encounter in clinics without a laboratory, rather than near-POC which still require minimal laboratory infrastructure and additional time before results are available. Testing for certain HPV genotypes can identify disproportionately more cervical intraepithelial neoplasia 2+ lesions (CIN2+; the precursor to cervical cancer) than HPV infections alone (7). For instance, a 4-target test in Africa (with HPV genotypes 16, 18, 35, and 45) would identify 26% of all HPV infections but capture 56% of CIN2+ cases (Figure 1)(1). While a limited-genotype test will identify fewer cases compared to a full-genotype panel under ideal screening conditions (i.e., equal coverage and perfect retention), there may be a use-case for such a test where access is poor or loss to follow-up is high(8). True POC limited-genotype HPV tests have only recently become commercially available; further development will be guided by a recently published target product profile(9).

**Figure 1.**
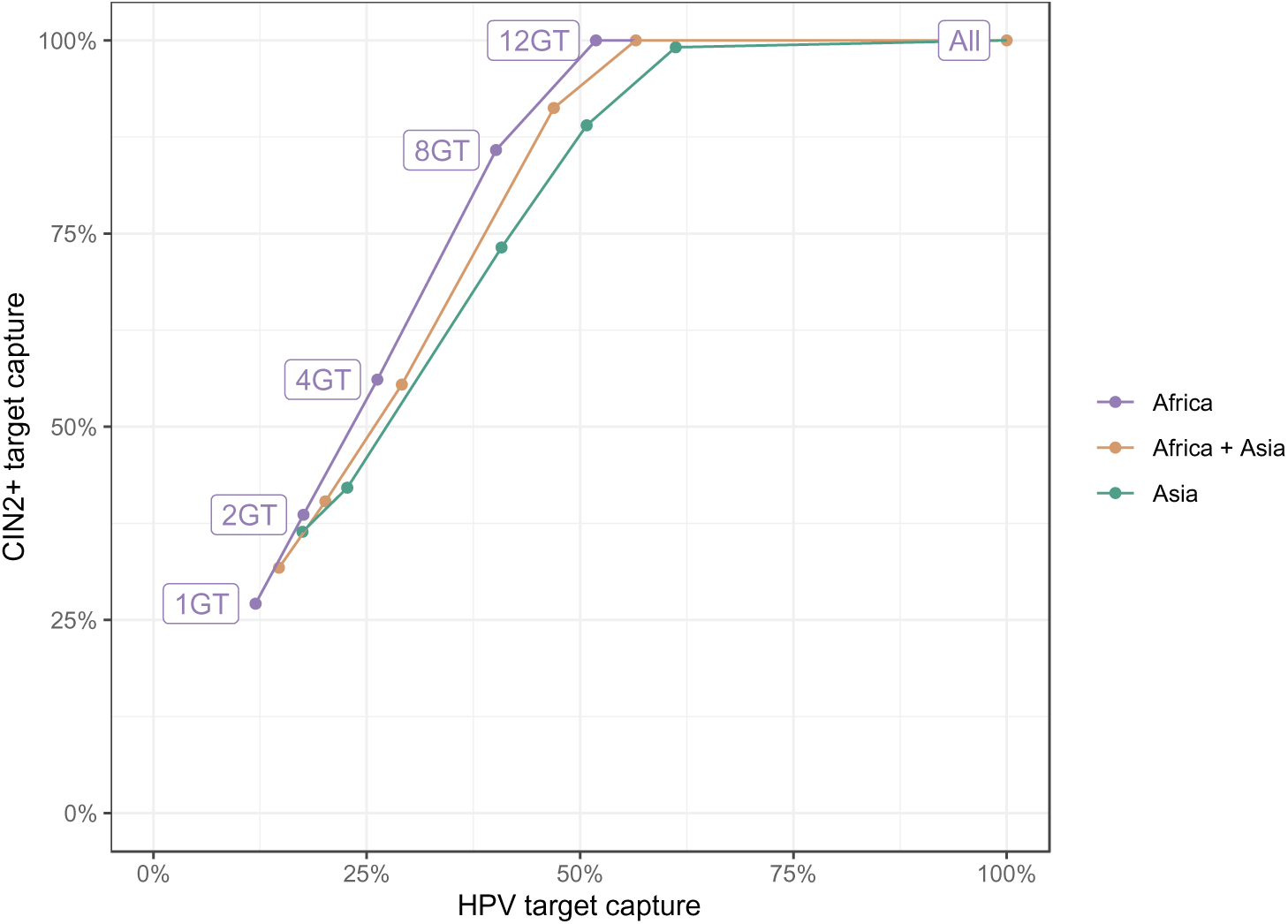
HPV and CIN2+ target capture for limited-genotype tests. Relationship between the percent of HPV infections and cases of cervical intraepithelial neoplasia 2+ (CIN2+) attributable to sets of limited HPV genotype targets (i.e., HPV and CIN2+ target capture, respectively). The optimal genotype target (GT) combinations were selected based on rank prevalence of each genotype among cervical cancer cases for the African and Asian regions separately and combined to demonstrate geographic variability in target capture. The included genotypes and target captures for 2, 4, 8, and 12 target tests in Africa are included in Table 1, as derived from data in (1).

**Table 1.**
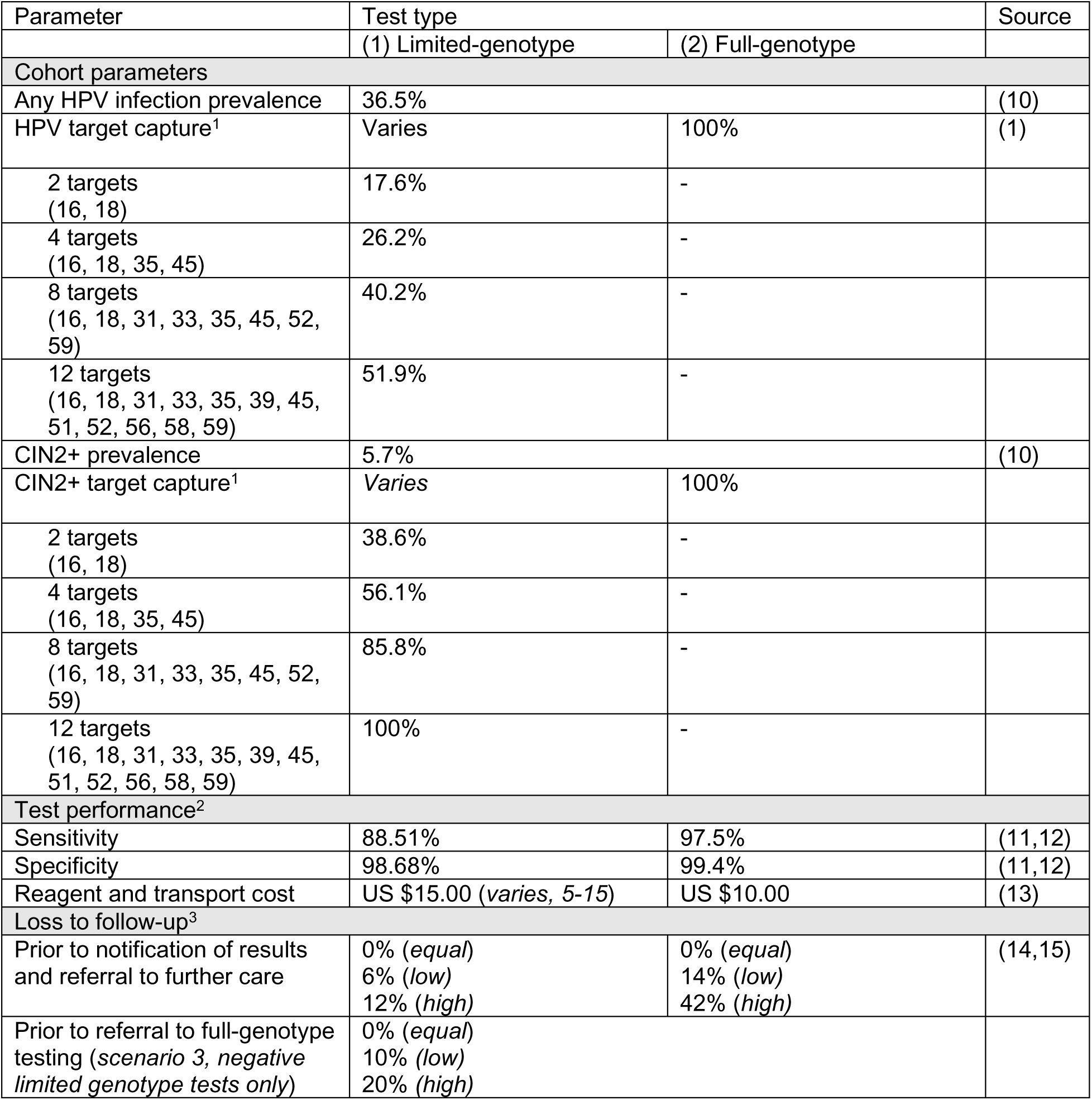
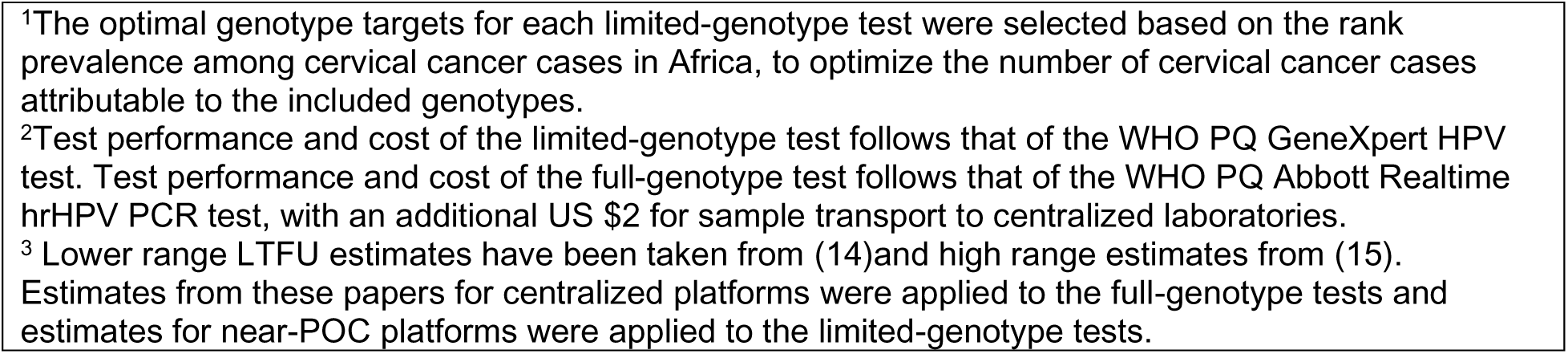
Model parameters in baseline scenarios; various sensitivity analyses are presented.

| Parameter | Test type |  | Source |
| --- | --- | --- | --- |
|  | (1) Limited-genotype | (2) Full-genotype |  |
| Cohort parameters |  |  |  |
| Any HPV infection prevalence | 36.5% |  | (10) |
| HPV target capture<sup>1</sup> | Varies | 100% | (1) |
| 2 targets<br>(16, 18) | 17.6% | - |  |
| 4 targets<br>(16, 18, 35, 45) | 26.2% | - |  |
| 8 targets<br>(16, 18, 31, 33, 35, 45, 52, 59) | 40.2% | - |  |
| 12 targets<br>(16, 18, 31, 33, 35, 39, 45, 51, 52, 56, 58, 59) | 51.9% | - |  |
| CIN2+ prevalence | 5.7% |  | (10) |
| CIN2+ target capture<sup>1</sup> | Varies | 100% |  |
| 2 targets<br>(16, 18) | 38.6% | - |  |
| 4 targets<br>(16, 18, 35, 45) | 56.1% | - |  |
| 8 targets<br>(16, 18, 31, 33, 35, 45, 52, 59) | 85.8% | - |  |
| 12 targets<br>(16, 18, 31, 33, 35, 39, 45, 51, 52, 56, 58, 59) | 100% | - |  |
| Test performance<sup>2</sup> |  |  |  |
| Sensitivity | 88.51% | 97.5% | (11,12) |
| Specificity | 98.68% | 99.4% | (11,12) |
| Reagent and transport cost | US \$15.00 (*varies, 5-15*) | US \$10.00 | (13) |
| Loss to follow-up<sup>3</sup> |  |  |  |
| Prior to notification of results and referral to further care | 0% (*equal*)<br>6% (*low*)<br>12% (*high*) | 0% (*equal*)<br>14% (*low*)<br>42% (*high*) | (14,15) |
| Prior to referral to full-genotype testing (*scenario 3, negative limited genotype tests only*) | 0% (*equal*)<br>10% (*low*)<br>20% (*high*) |  |  |

<sup>1</sup>The optimal genotype targets for each limited-genotype test were selected based on the rank prevalence among cervical cancer cases in Africa, to optimize the number of cervical cancer cases attributable to the included genotypes.
<sup>2</sup>Test performance and cost of the limited-genotype test follows that of the WHO PQ GeneXpert HPV test. Test performance and cost of the full-genotype test follows that of the WHO PQ Abbott Realtime hrHPV PCR test, with an additional US \$2 for sample transport to centralized laboratories.
<sup>3</sup> Lower range LTFU estimates have been taken from (14) and high range estimates from (15). Estimates from these papers for centralized platforms were applied to the full-genotype tests and estimates for near-POC platforms were applied to the limited-genotype tests.

Here, we model the trade-offs between number of HPV genotype targets, test cost, retention, and coverage to inform target product profiles for the development of new HPV testing technologies for low and middle-income countries. The tradeoff is between a hypothetical POC limited genotype test with better follow-up and access but fewer cases of precancer identified, versus a centralized full genotype HPV test with lower follow-up and less access but more cases of precancer identified. Our work provides guidance to product developers and manufacturers on the required test performance, target price, and conditions for limited-genotype tests to be an attractive alternative to current diagnostics.

## Methods

We developed a simplified diagnostic screening model to assess the performance of limited-genotype tests in identifying cases of CIN2+ against conventional screening for all high-risk HPV genotypes (Figure 2).

**Figure 2.**
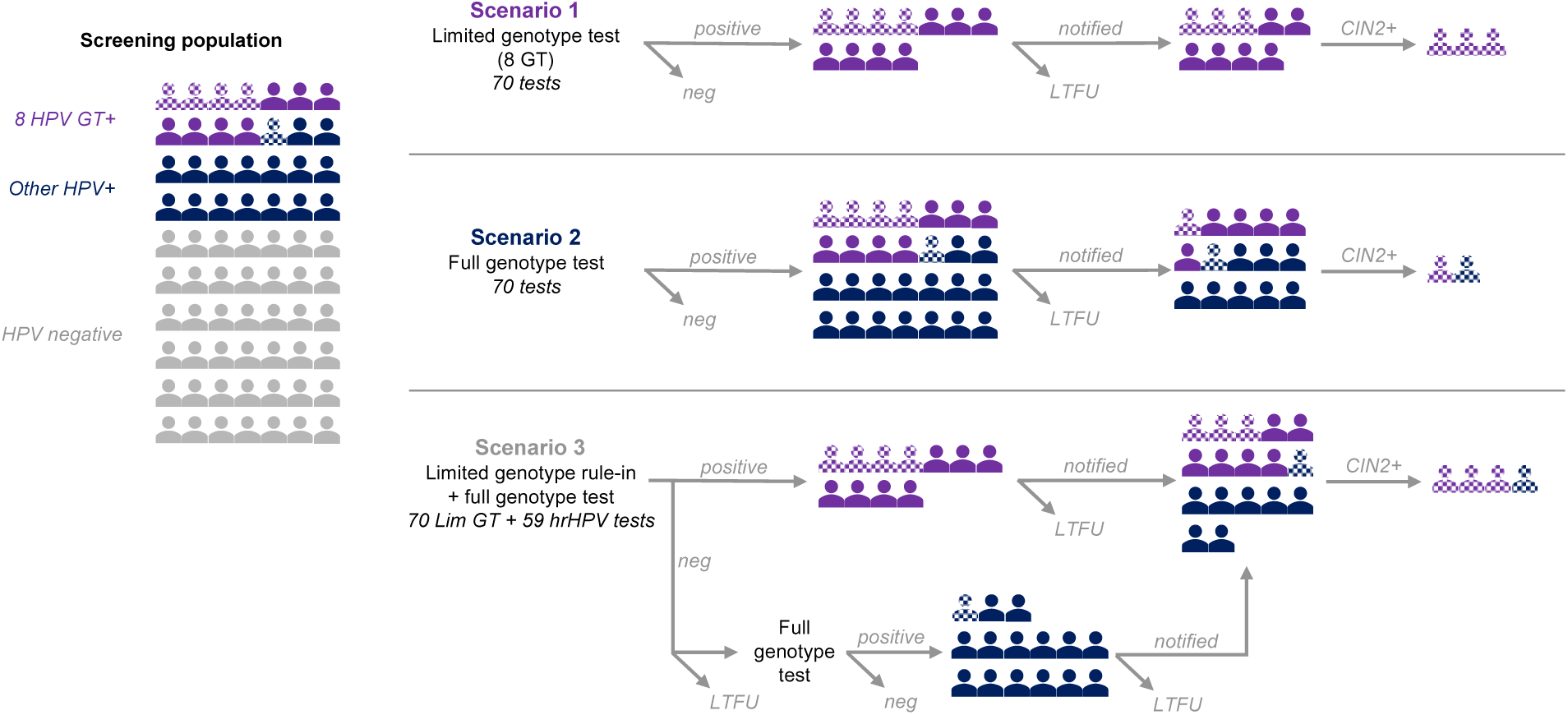
HPV screening model diagram. Schematic of HPV screening pathways with (1) a point-of-care limited genotype test, here with 8 genotype targets (GT); (2) a molecular test of all genotypes (full genotype test) performed at a centralized laboratory; and (3) a rule-in limited genotype target test followed by a full genotype test for those who screened negative, where loss to follow-up (LTFU) can occur at multiple steps. Colors represent HPV infection status and shaded figures have cervical intraepithelial neoplasia grade 2+ (CIN2+). Note that in this schematic, we assume all tests are perfectly sensitive and specific; in the model, imperfect sensitivity and specificity give rise to false positive and false negative test results. Schematic represents an example population of n=70; actual model parameters are presented in Table 1.

### Model structure

We simulated a cohort of 1,000 women accessing cervical cancer screening services at healthcare facilities. The cohort was divided by HPV infection and CIN2+ disease status, given the underlying total and genotype-specific prevalence of each. In baseline models, all sought care and received screening services, comprised of limited genotype target (“limited-genotype”) HPV molecular tests or comprehensive molecular tests of all hrHPV genotypes (“full-genotype”). Screening coverage was varied by test type in sensitivity analyses.

Women who underwent screening were assigned either a positive or negative test result, based on infection status and the diagnostic accuracy of the available test. For limited-genotype tests, sensitivity was defined as the probability of correctly identifying individuals with an infection of one or more included HPV genotype, and specificity was defined as the probability of correctly identifying individuals without any infection of an included HPV genotype (but possibly with infection by other HPV genotypes). We defined the proportion of all HPV infections and CIN2+ cases attributable to genotypes included in the limited-genotype test as the HPV target capture and CIN2+ target capture, respectively. We only considered detection of hrHPV infections in this model and assumed that all CIN2+ cases were attributable to hrHPV infection, such that HPV and CIN2+ target capture for the full-genotype test are both 1.

### Scenario parameterization and comparison

We modelled three scenarios: (1) all women receiving a limited-genotype HPV test, where the CIN2+ target capture is varied to reflect different numbers of included genotype targets (“**limited-genotype”)**; (2) all women receiving a full-genotype test (“**full-genotype**”); and (3) all women receiving a limited-genotype HPV test, followed by a full-genotype test for those screening negative (“**limited + full-genotype**”) (Figure 2). This last scenario arose out of discussions around the perceived inequity and negative consequences of screening only for a limited set of genotypes without follow-up testing.

To account for loss to follow-up prior to notification and linkage to further testing and treatment, our primary outcome was the number of women with CIN2+ who correctly tested positive and were notified of their disease status for each testing strategy. We assumed that women who screened positive and were notified of their result were referred for treatment or appropriate confirmatory testing. As we were interested in comparing diagnostic pathways across otherwise hypothetically equivalent populations and healthcare facilities, we did not explicitly model treatment referral and receipt following notification. The probabilities of loss to follow-up following screening and, when varied, of accessing screening services were independent of disease status and test result.

In scenario 3, loss to follow-up could occur: (1) following limited-genotype testing, prior to notification; (2) during patient or sample referral to full-genotype testing, following a negative limited genotype result; and (3) following full-genotype testing, prior to notification. In practice, we expect this kind of multistage screening strategy would involve simultaneous collection of two samples at the time of limited-genotype testing, to minimize required clinic visits for follow-up testing, such that loss to follow-up between limited-genotype and full-genotype testing (point 2) should be due primarily to gaps in the sample referral system.

In all scenarios, we estimated the total screening costs as the sum of all women tested with each test type times the cost of the test used, including costs of test reagents and transport if necessary. We did not include costs of personnel or patient time in our estimation. Since two samples are collected upfront in scenario 3, an additional clinical appointment is not required, and additional costs are not incurred. Key model outcomes for each scenario were the proportion of all CIN2+ cases identified and notified (the CIN2+ cases correctly identified by the test and not lost to follow-up) and the total testing cost per CIN2+ case identified for each scenario. To assess whether scenario 1 and 3 were non-inferior relative to scenario 2 (the assumed ‘standard of care’ or gold-standard) we compared the average cost effectiveness ratio or the cost per outcome of the scenario to the cost per outcome calculated for scenario 2. In addition, we determine the ‘maximum allowable cost’ of a limited-genotype test, with and without full-genotype testing for negatives, in order to match the cost of full-genotype screening across a range of conditions (CIN2+ target capture, full-genotype loss to follow-up).

Key model parameter values, including HPV prevalence, CIN2+ prevalence, HPV and CIN2+ target capture, test performance, and cost are included in Table 1. Notably, we assumed that limited-genotype tests could be performed at the point of care, whilst conventional full-genotype testing was performed offsite at centralized laboratories, as per current use of full-genotype HPV testing. Therefore, loss to follow-up was assumed to be lower following limited-genotype testing, where notification and linkage to care could be provided within the same clinical encounter, compared to full-genotype testing where loss to follow-up was assumed to be higher due to the need for sample transport to centralized laboratories and for a second clinical encounter for notification and linkage to care. All modeling and analysis was conducted in R version 4.3.0.

## Results

### Limited-genotype tests required sufficient target capture to compete with case identification of full-genotype screening

Under reasonable assumptions of test performance and low assumed loss to follow-up (6% limited-genotype, 14% full-genotype) (Table 1), screening with a full-genotype test identified and referred 83.9% of CIN2+ cases to further care (Figure 3A, Table S1). Regardless of target capture, full-genotype screening identified more cases of CIN2+ than limited-genotype tests alone. When the model was parametrized with high assumed loss to follow-up (12% limited-genotype, 42% full-genotype), limited-genotype tests with ≥72% CIN2+ target capture (between 4 and 8 genotype targets) were competitive with full-genotype testing (Figure S1A).

**Figure 3.**
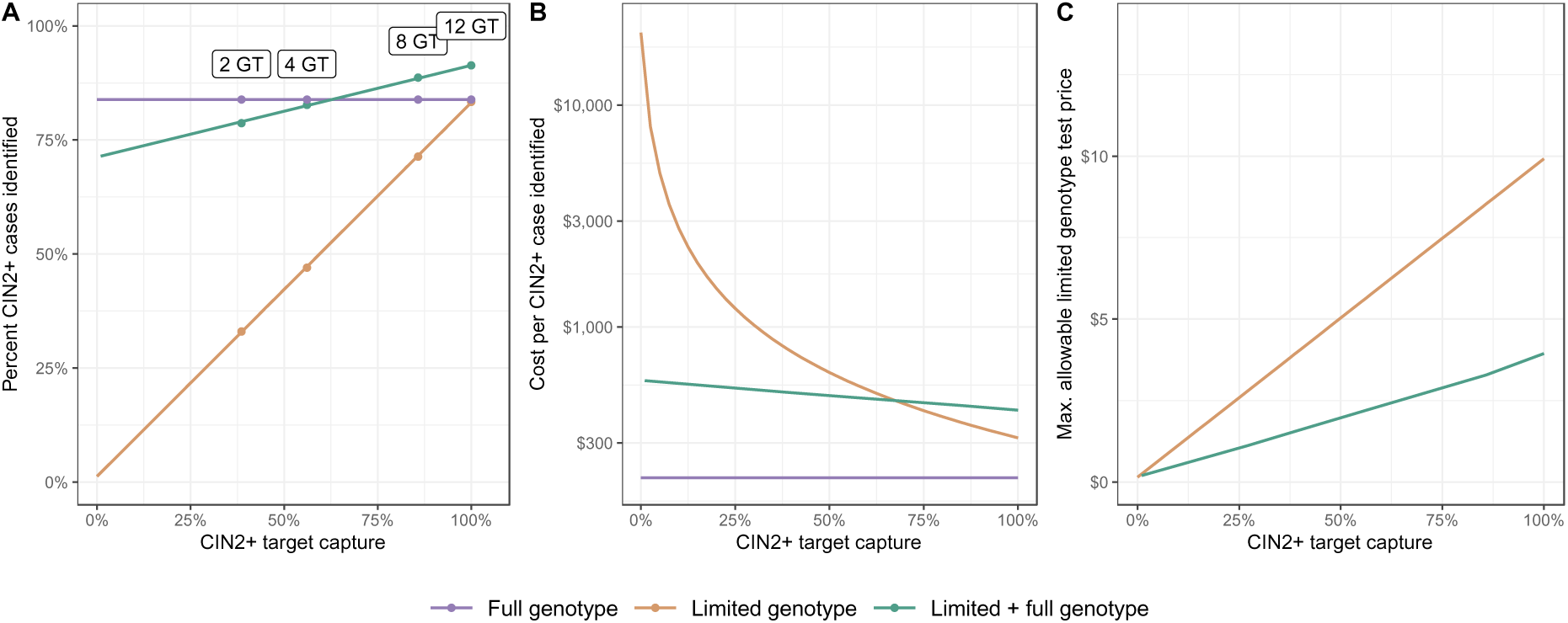
HPV screening outcomes with limited-genotype and full-genotype tests. (A) CIN2+ cases captured and (B) cost per case captured screening with a US $10 full-genotype test (scenario 2), a US $15 limited genotype test (scenario 1) and limited genotype test followed by full-genotype testing (scenario 3), and (C) the maximum allowable cost of a limited-genotype test, with and without full-genotype testing for negatives, in order to match the cost of full-genotype screening across a range of CIN2+ target capture values for the limited-genotype test and with low loss to follow-up. The labels in panel A mark the CIN2+ target capture for optimal genotype (GT) combinations in Africa (e,g, 2 GT).

Under low assumed loss to follow-up, when limited-genotype tests with ≥62% CIN2+ target capture (>4 genotype targets) were used as a rule-in test with referral to full-genotype testing for women screening negative (scenario 3), more cases of CIN2+ were identified than with full-genotype screening alone. However, under high assumed loss to follow-up, a limited-genotype rule-in test with just 38% CIN2+ target capture (2 genotype targets) could identify as many CIN2+ cases as full-genotype screening (Figure S1A). Though the addition of full-genotype testing always improved case identification compared to limited-genotype testing alone, the relative benefits were reduced when loss to follow-up was high and when target capture was high.

### Limited-genotype test price was a key determinant of its viability

Test costs for limited-genotype tests will be a barrier to implementation relative to existing molecular diagnostic platforms providing full-genotype testing. While the cost per CIN2+ case identified decreased as target capture increased, there was no scenario where a US $15 limited-genotype test, alone or as a rule-in test, was less costly than full-genotype testing (Figure 3B, Figure S1B). When loss to follow-up was low, limited-genotype tests needed to cost less than full-genotype tests (US $10) to be cost-equivalent; for example, an 8-target limited-genotype test (86% CIN2+ target capture) needed to cost less than US $8.50 to match the cost per case identified with full-genotype screening alone (Figure 3C). The maximum allowable limited-genotype test price increased with greater target capture and with greater reductions in loss to follow-up with POC testing (Figure S1C), such that when loss to follow-up was high, limited-genotype tests with sufficient target capture (≥70%) used without full-genotype testing for negatives could cost more than US $10.

When used as a rule-in diagnostic, the allowable cost of the limited-genotype test was substantially lower than limited-genotype screening alone (less than US $5 at all target capture values), to account for the additional costs of full-genotype testing for negatives (Figure 3C). The additional case identification benefits of rule-in testing, however, led to a lower cost per case than limited-genotype testing alone when target capture was low (Figure 3B). Limited-genotype screening followed by full-genotype testing became more expensive than limited-genotype screening alone once sufficient CIN2+ target capture was reached (≥66% or >4 targets in the low loss to follow-up scenario; 57% or 4 targets in the high loss to follow-up scenario), thereby reducing the number of additional cases identified with full-genotype testing for negatives (Figure 3B, Figure S1B).

These relationships were reflected in the pairwise incremental cost effectiveness ratio of each limited-genotype testing scenario to the full-genotype scenario (**Error! Reference source not found.**).

### Limited-genotype test requirements depend on loss to follow-up under centralized testing

The assumed retention following full-genotype testing through centralized laboratories had substantial influence over the conditions in which limited-genotype testing was non-inferior. The required CIN2+ target capture for a limited-genotype test alone to be competitive with full-genotype tests in terms of case identification decreased linearly with increasing loss to follow-up after full-genotype testing (Figure 4A). An 8-target (86% CIN2+ target capture) limited-genotype test with 6% LTFU identified more cases than full-genotype testing with ≥19.8% loss to follow-up. At sufficiently high (≥63.4%) loss to follow-up after full-genotype testing, even a 2-target (38.6% CIN2+ target capture) limited-genotype test with low attrition could identify more cases of CIN2+.

**Figure 4.**
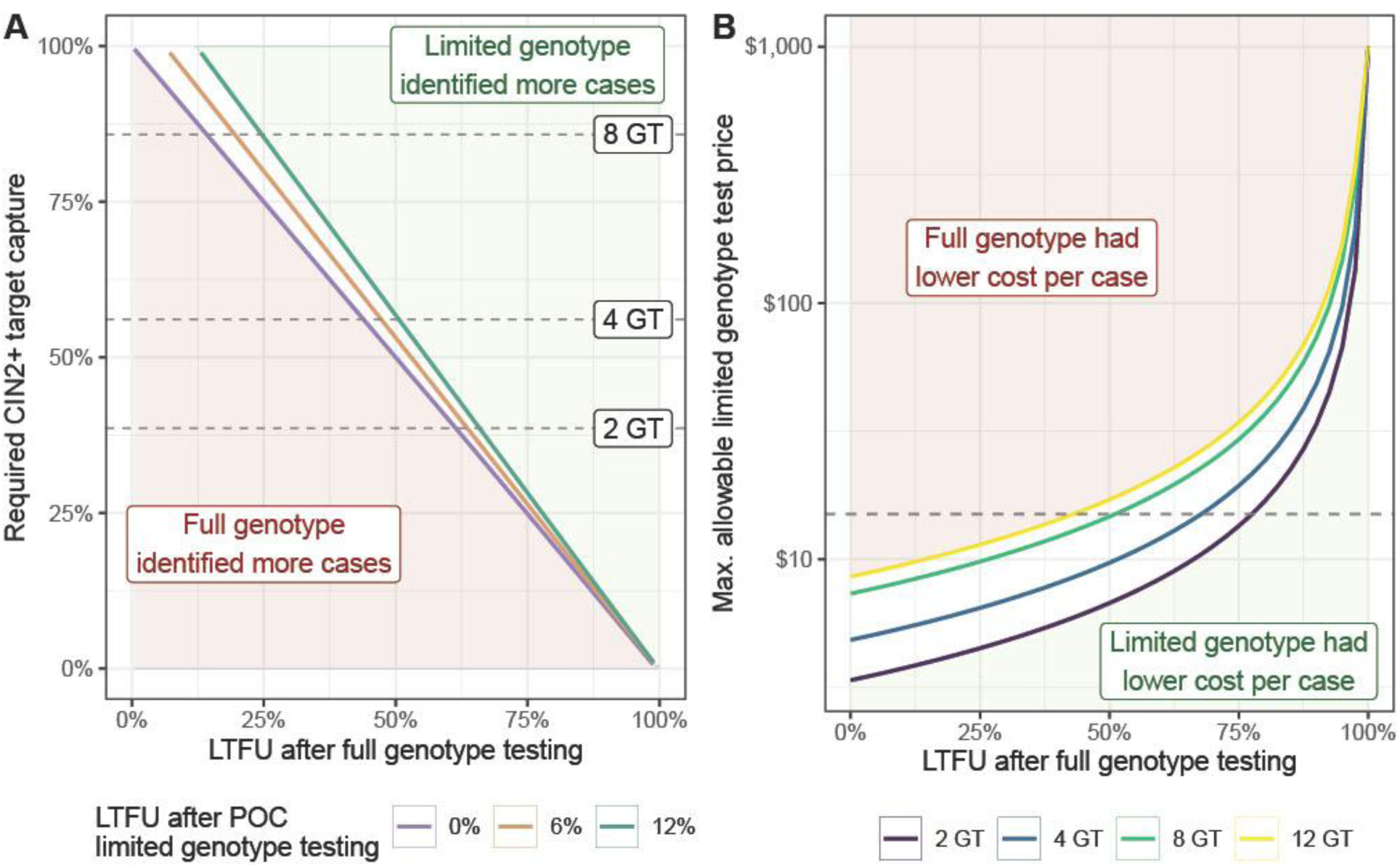
Impact of loss to follow-up following centralized full HPV genotype tests. The influence of assumed loss to follow-up (LTFU) following full-genotype testing on (A) minimum required CIN2+ target capture of limited-genotype tests to be non-inferior to full-genotype testing in terms of the proportion of CIN2+ cases identified and (B) the maximum allowable limited-genotype test price (with 6% LTFU) to be cost-equivalent to full-genotype testing. Panel (A) shows the required CIN2+ target capture for 3 values of LTFU following POC limited-genotype testing, and panel (B) shows the required test price for the optimal 2, 4, 8, and 12-target limited-genotype tests in Africa. The dashed lines in panel (A) indicate CIN2+ target capture with 2, 4, and 8 targets, and the dashed line in panel (B) represents a limited-genotype test price of US $15. Red shaded regions show where full genotype cases outperformed limited genotype tests (i.e., identified more cases or had lower cost per case identified) for all values of POC loss to follow-up or genotype target, and green regions show where limited genotype tests outperformed full genotype tests. Unshaded regions show where differences in POC loss to follow-up or target capture would influence whether full genotype or limited genotype tests would have better performance.

Limited-genotype tests with a higher cost and lower target capture could be competitive with full-genotype testing if loss to follow-up was high. While limited-genotype tests had to cost less than US $10 when loss to follow-up was low (Figure 3C), the maximum allowable test price increased sharply with higher loss to follow-up (Figure 4B), such that an 8-target and US $15 limited-genotype test was cost-equivalent to full-genotype screening when loss to follow-up was >51% with full-genotype testing. The same relationships were observed when rule-in limited-genotype tests were combined with full-genotype testing for negatives (scenario 3), though again the maximum allowable price was overall lower than for limited-genotype tests offered alone (Figure S2).

### Increased screening coverage can outweigh reduced sensitivity of limited-genotype tests

A significant potential advantage to limited-genotype tests is the ability to expand screening coverage to areas with poor or no access to centralized laboratories or sample referral systems for full-genotype testing. There were direct trade-offs between potential increases to screening coverage through limited-genotype tests and the needed CIN2+ target capture to compete with full-genotype testing (Figure 5). For example, an 8-target limited-genotype test necessitated only a 5% increase in screening coverage with low loss to follow-up, or as much as a 45% increase for a 2-target limited-genotype test, to identify the same number of CIN2+ cases as full-genotype screening. The required changes to screening coverage increased as loss to follow-up decreased, and therefore the relative difference in retention for centralized full-genotype and POC limited-genotype testing decreased (Figure S3). When loss to follow-up was equal between limited-genotype and full-genotype testing (e.g., if limited-genotype tests were also conducted at centralized laboratories through a sample referral system), screening coverage needed to increase by >8% for an 8-target limited-genotype test or >54% for a 2-target limited-genotype test.

**Figure 5.**
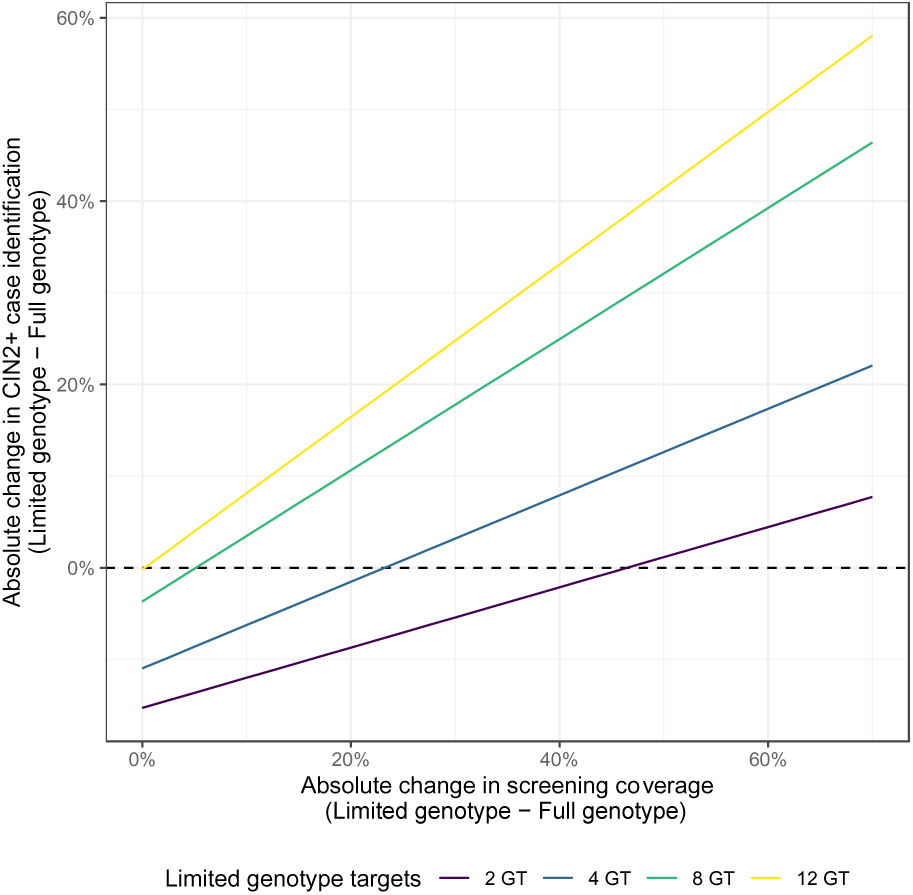
Trade-offs between screening coverage and target capture of CIN2+ cases using limited-genotype versus full-genotype screening. The figure illustrates the absolute increase in the percentage of all CIN2+ cases detected when screening coverage with limited-genotype tests is between 0% and 70% higher than with full-genotype tests, assuming a baseline coverage of 30%. This comparison is shown under scenarios of low loss to follow-up (6% for point-of-care and 14% for centralized screening), highlighting the potential to capture a greater number of CIN2+ cases by expanding screening coverage with limited-genotype tests.

The relationships between screening coverage, target capture, and loss to follow-up were similar when full-genotype testing was conducted for those negative on the limited-genotype test (scenario 3). Due to the additional case identification with full-genotype testing, however, the required increases to screening coverage to be equivalent to full-genotype testing alone were overall lower than for limited-genotype tests alone, particularly for limited-genotype tests with few targets (Figure S4).

### Disease prevalence had limited impact on screening performance

The underlying burden of HPV and CIN2+ influenced the costs of each testing strategy. When HPV and CIN2+ prevalence were high, fewer tests were needed to identify a CIN2+ case, resulting in lower costs per case identified for all scenarios (Figure S5A). The relative impacts, however, were the same for limited-genotype and full-genotype testing alone, such that the allowable cost of a limited-genotype test without referral to full-genotype testing for negatives was unaffected by disease prevalence (Figure S5B).

The compounding effect of increased case identification with limited-genotype tests and therefore fewer referrals to full-genotype testing led to relatively greater reductions in the cost per case identified with rule-in limited-genotype tests followed by full-genotype testing, compared to limited-genotype or full-genotype testing alone, when disease prevalence was high. However, the magnitude of the effect was small. This resulted in a higher maximum allowable cost of limited-genotype tests followed by full-genotype testing compared to the baseline prevalence scenario (Figure S5B). These relationships were reversed when prevalence was low: cost per case identified increased for all scenarios, especially in the case of scenario 3, and the allowable price of limited-genotype tests followed by full-genotype testing decreased (Figure S6)

## Discussion

Limited-genotype target tests for HPV infection can be an important tool in achieving cervical cancer elimination targets, particularly where access to centralized testing is currently poor. While previous work modelled tradeoffs between test sensitivity, coverage, retention, and costs of molecular HPV testing versus VIA(8), our model explicitly considered the implications of adopting limited-genotype HPV tests at point-of-care within a cervical cancer screening program.

Using a simplified model of a once-off screening, we demonstrated that, when centralized full-genotype screening is performing optimally (e.g., if a full-genotype test can be offered as a true POC test with high retention(16)), POC limited-genotype screening alone will identify fewer cases of CIN2+, but may be less expensive in identifying a CIN2+ case with a sufficiently low test price. However, when loss to follow-up after full-genotype testing is high, or when access to testing can be expanded through limited-genotype screening at facilities with no sample or patient referral systems in place, limited-genotype tests can be highly competitive even with a reduced number of targets(17).

The exact target capture needed is contingent on the assumed screening coverage and loss to follow-up after full-genotype testing; higher loss to follow-up with full-genotype testing and greater access to limited-genotype tests resulted in greater tolerance for reduced target capture. Generally, based on our modelling results, a limited-genotype test with ≥8 targets will be non-inferior in terms of both case identification with modest increases (≥5% with low loss to follow-up) in screening coverage or sufficiently high loss to follow-up (≥19.8%) after full-genotype testing. An 8-target test would also be practical for widespread use, given variance in regional genotype distributions and the resulting difficulty in selecting universally ideal genotype combinations with fewer targets. In certain contexts (e.g., when full-genotype loss to follow-up is greater than 60%, as observed for viral load results in 7 African countries(15)), even a 2-target, HPV-16/18 limited-genotype test could capture a higher proportion of CIN2+ cases than full-genotype screening. However, in these contexts with high loss to follow-up, the adoption of limited-genotype tests would need to be weighed up against other interventions such as digital solutions and programmatic improvements (such as community healthcare worker programs) that can improve retention using centralized testing.

Reducing the cost of limited-genotype tests will be critical for the feasibility and cost-effectiveness of their adoption relative to existing full-genotype tests. At a test cost of US $15 (close to currently available POC technology for a full-genotype test), there was no scenario at which a limited-genotype test with less than 100% target capture had a lower cost per case identified than full-genotype testing, under reasonable assumptions of loss to follow-up. The required price for limited-genotype tests to be cost-equivalent to full-genotype testing depends on the target capture and assumed loss to follow-up. With low loss to follow-up, a limited-genotype test with sufficient target capture (≥8 targets) could cost up to US $8.50 to be cost-equivalent to a full-genotype scenario on a cost per case identified basis. Relatedly, reducing the test price allows limited-genotype screening to be cost-competitive across a wider range of target capture levels and levels of loss to follow-up of centralized full-genotype testing. The recently published target product profile for an HPV point of care test for a limited number of genotypes (8 targets) has a cost per reportable result an amount of $8, with a preferred amount of $3 (inclusive of consumables and collection devices) (9). This suggests that, following consultations with experts, it is considered feasible to lower the test price further making a limited genotype test even more cost-competitive.

Even where limited-genotype screening may identify more cases overall than full-genotype screening, there would still be a proportion of the population with HPV infection, and thus risk of severe disease, that would not be captured in the reduced genotype targets. Using limited-genotype testing as a rule-in diagnostic with full-genotype testing for all women with negative results is one way to improve and address inequities in case identification. The addition of a full-genotype test is costly, though, and while such a rule-in scenario with a 4-target limited-genotype test can be just as effective as full-genotype screening with low loss to follow-up, it will be more expensive unless the limited-genotype tests cost less than US $2.20. As with limited-genotype screening alone, lower CIN2+ capture and greater test price can be tolerated if there is high loss to follow-up after full-genotype screening alone or increased screening coverage. Adding a rule-in test becomes less attractive than limited-genotype screening alone when the limited-genotype test alone has sufficient target capture (≥6 targets with low loss to follow-up), such that the follow-on full-genotype testing captures relatively few additional cases.

There are a number of important considerations and limitations to our modeling approach. First, in using the CIN2+ target capture as our measure of genotype coverage, we have not modeled which HPV genotypes should be included in a limited-genotype test. The heterogenous distribution of HPV genotypes globally means a fixed set of genotypes will have different CIN2+ target captures in different contexts, leading to different optimal combinations of HPV genotypes. This effect would be particularly noticeable with limited-genotype tests with 4 targets or fewer and may necessitate designing regionally-specific limited-genotype tests, which presents logistical and regulatory challenges. With 8 or more genotypes, CIN2+ target capture will be more consistent globally and likely sufficient in all contexts to be competitive with full-genotype testing. Our model also implicitly assumes the target capture value would remain constant; in reality, genotype-specific screening and scale-up of HPV vaccination could exert selective pressure resulting in changing genotype distributions over time, which could complicate the development and deployment of limited-genotype tests and necessitate the re-evaluation of the target choice in the assay at a regular interval.

Our goal was to identify the conditions under which limited-genotype tests could be used in place of full-genotype tests with equivalent or non-inferior outcomes. As such, our model considers only a once-off screening event with intermediate outcomes (number CIN2+ cases identified and notified), after which downstream benefits and costs of each screening program are assumed to be equivalent. Utilizing a longitudinal screening model and cost-utility analysis would also be more suitable for depicting the lifetime costs and benefits of a cervical cancer screening program with either test, particularly where triage and clinical management following either test may vary(18). This approach would also allow for comparisons to competing health interventions targeting different diseases and for exploration of alternate cervical cancer screening intervals, another possible lever to improve the performance and cost of screening programs with limited-genotype tests.

The impact of any molecular test will be dependent on successful referral to further evaluation and treatment, either in screen-and-treat or screen-triage-and-treat programs. We assumed, for the purposes of evaluating limited-genotype tests as an alternative to full-genotype screening, that the care cascade following notification was identical for all modeled scenarios. Expanding diagnostic access through POC limited-genotype tests without concomitant expansion of VIA and treatment capacity, though, will attenuate impact and cost-effectiveness of limited-genotype screening. Our model also considered the positive identification of true CIN2+ cases as the primary outcome of interest, to ensure maximum impact and equity in screening programs. This ignores the costs associated with increased treatment of women without HPV infection or CIN2+ that might arise from the lower specificity of POC tests. We note, however, that restricting testing to a limited set of genotypes has been shown to improve the specificity and positive predictive value of HPV screening for identifying cases of CIN2+(7).

Here, we have conducted a non-inferiority analysis with full-genotype screening as the comparator and cost-effectiveness threshold for our analysis. If the cost per CIN2+ case identified was greater with limited-genotype screening, we concluded that limited-genotype screening was a less efficient strategy than full-genotype screening (alternatively, it could be interpreted that for the same budget, a policymaker could buy the identification of fewer CIN2+ cases). Using the comparator as a cost-effective threshold is a more stringent threshold than other commonly-used thresholds(19), but allows for direct assessment of when a limited-genotype test would be non-inferior to the gold-standard full-genotype test. We also considered only costs associated with test reagent and transport costs, rather than a full costing of each screening program. Again, our objective was to determine the price of a POC limited-genotype assay that would be cost-equivalent in place of an offsite full-genotype assay. We believe this price point is more informative to manufacturers who would be developing the test and more intuitive across different contexts where the price would be consistent but the other cost elements more variable.

Lastly, we have considered limited-genotype screening as an alternative to full-genotype screening; in reality, the two will likely be scaled together alongside other interventions that are likely to improve access, such as self-collection of specimens. While our results provide guidance for the contexts in which one test may be favored over the other and the trade-offs between test sensitivity, access, retention, and cost, further work is needed to determine the optimal implementation strategy for limited-genotype tests within national programs.

### Conclusions

Under the specified conditions of coverage and loss to follow-up, our analysis supports the use of an 8-target, limited genotype test priced at less than US $8.50 for cost-equivalence with full-genotype screening. A 4-target limited genotype test can be effective as a rule-in test with referral to full-genotype screening, but must cost less than US$2.20 to be competitive with full-genotype screening. Our flexible, parsimonious modeling framework can help decision-makers identify the various conditions of test access, loss to follow-up, and cost under which the adoption of limited genotype screening can lead to better outcomes than centralized full-genotype screening alone. These insights can guide the development and implementation of limited genotype testing technologies to enhance screening access and support the goal of eliminating cervical cancer.

## Declarations

### Ethics approval and consent to participate

Not applicable.

### Consent for publication

Not applicable.

### Availability of data and materials

All data generated or analyzed during this study are included in this published article and its supplementary information files. The core model functions are available in the Zenodo repository (https://zenodo.org/doi/10.5281/zenodo.10127147).

### Competing interests

The authors declare that they have no competing interests.

### Funding

With support from *Canada’s* Department of Foreign Affairs, Trade and Development (*DFATD*)

### Authors’ contributions

KHG, SG, SK contributed to the conception and design of this work. KHG and SG performed analysis and wrote the first draft, and all authors contributed to the interpretation of results and revision of the manuscript. All authors have read and approved the submitted version.

## Acknowledgements

Not applicable.

## Notes

### Competing Interest Statement

The authors have declared no competing interest.

