## supplementary file for "Assessing feasibility and effectiveness of point-of-care limited HPV genotype tests in cervical cancer screening: a modelling study"

**Additional File 1:** Supplementary Tables and Figures

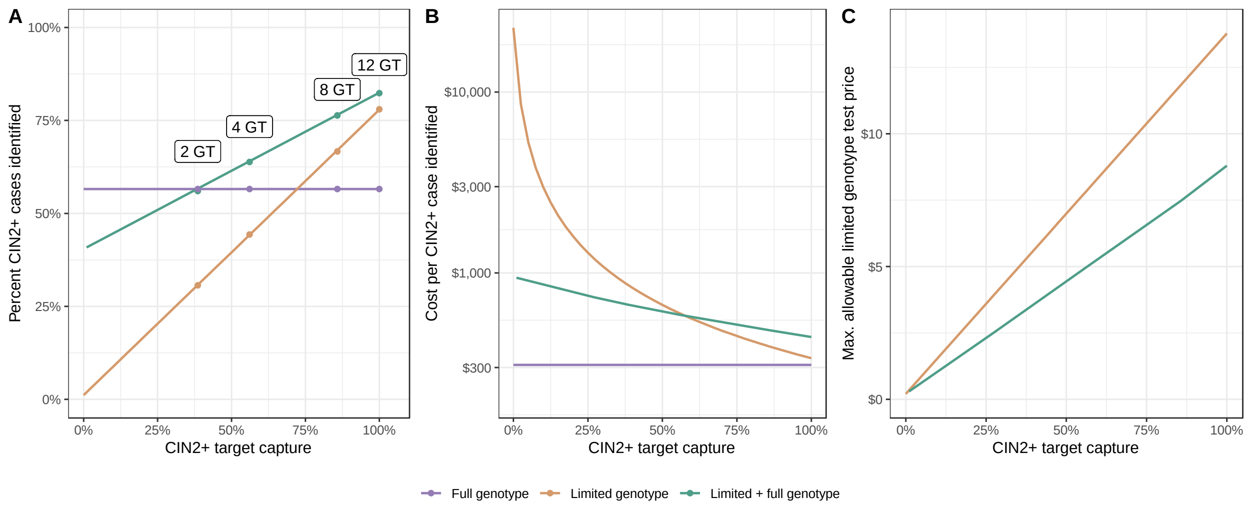


*Figure S1. (A) CIN2+ cases captured and (B) cost per case captured screening by full-genotype testing (scenario 1), limited-genotype screening (scenario 2) and rule-in limited-genotype screening followed by full-genotype testing (scenario 3), and (C) the maximum allowable cost of a limited-genotype test in order to match the cost of full-genotype screening, with and without full-genotype testing for negatives, across a range of CIN2+ target capture values for the limited-genotype test and with high loss to follow-up. Labels mark the CIN2+ target capture for optimal genotype combinations in Africa.*

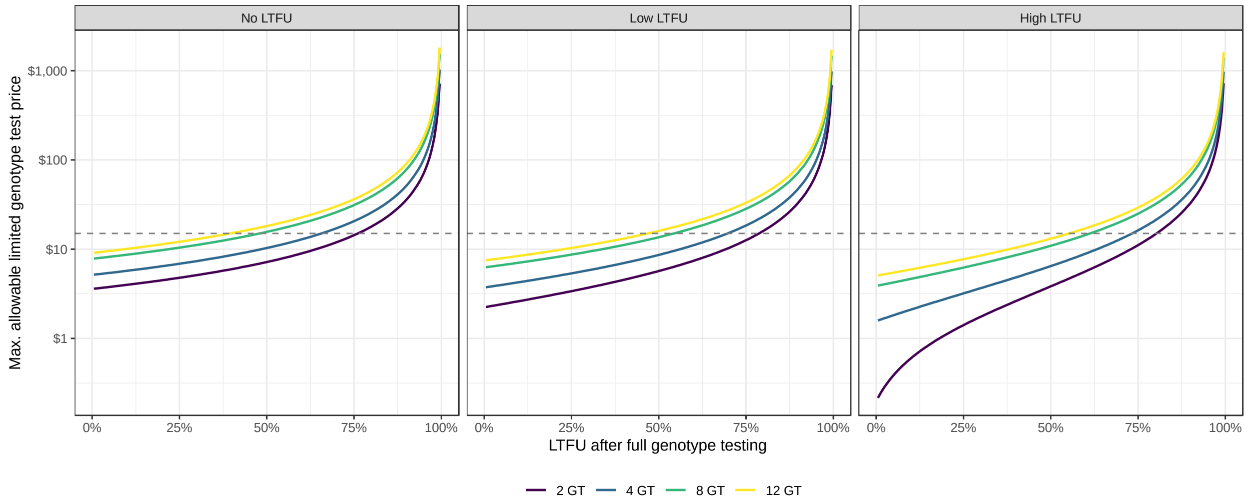


*Figure S2. The influence of assumed loss to follow-up (LTFU) following centralized full genotype testing on the maximum allowable price of rule-in limited-genotype tests when combined with full-genotype testing (scenario 3), in order to be cost-equivalent to full-genotype screening. Each line shows the required test price for the optimal 2, 4, 8, and 12-target limited-genotype tests in Africa, across three scenarios of loss to follow-up for limited-genotype testing.*

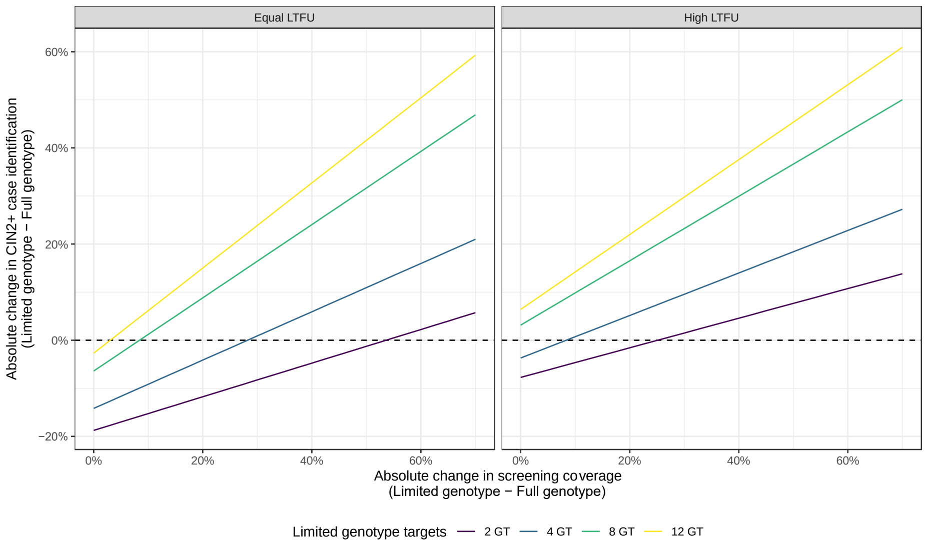


*Figure S3. The absolute increase in the percent of all CIN2+ cases positively identified through limited-genotype screening compared to full-genotype screening, when loss to follow-up is equal (0% both tests) or high (12% POC limited-genotype, 42% centralized full-genotype) and when screening coverage with limited-genotype tests between 0% to70% higher than with full-genotype tests, assuming a baseline coverage of 30%, highlighting the potential to capture a greater number of CIN2+ cases by expanding screening coverage with limited-genotype tests.*

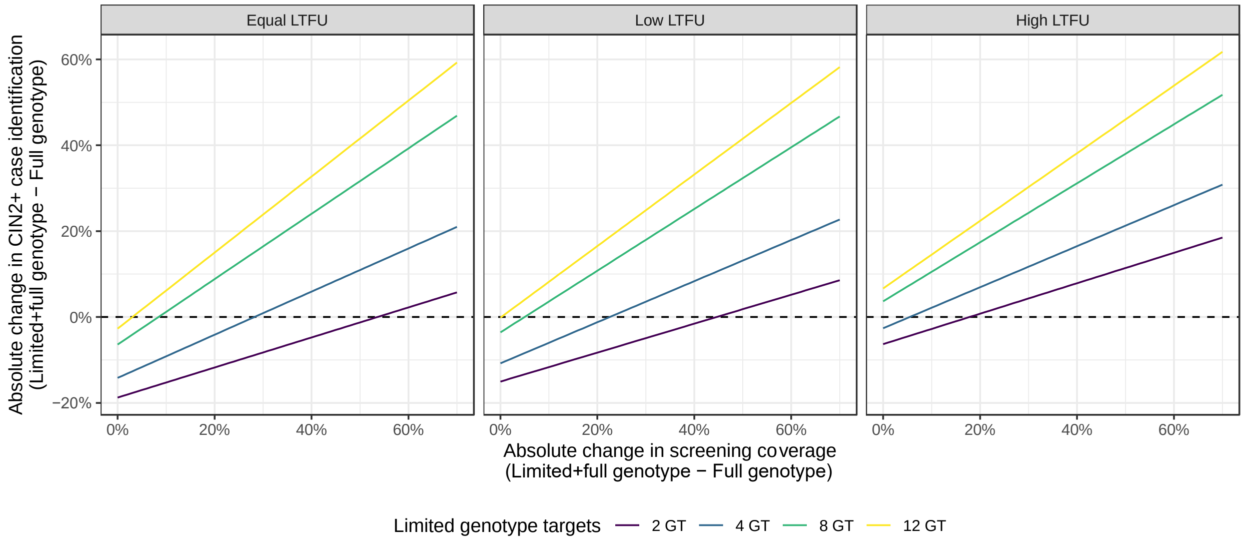


*Figure S4. The absolute increase in the percent of all CIN2+ cases positively identified through rule-in limited-genotype testing with full-genotype testing for those testing negative (scenario 3), compared to full-genotype testing (scenario 1) when loss to follow-up is equal (0%), low, or high and when screening coverage with limited-genotype tests is between 0% to70% higher than with full-genotype tests, assuming a baseline coverage of 30%, highlighting the potential to capture a greater number of CIN2+ cases by expanding screening coverage with limited-genotype tests.*

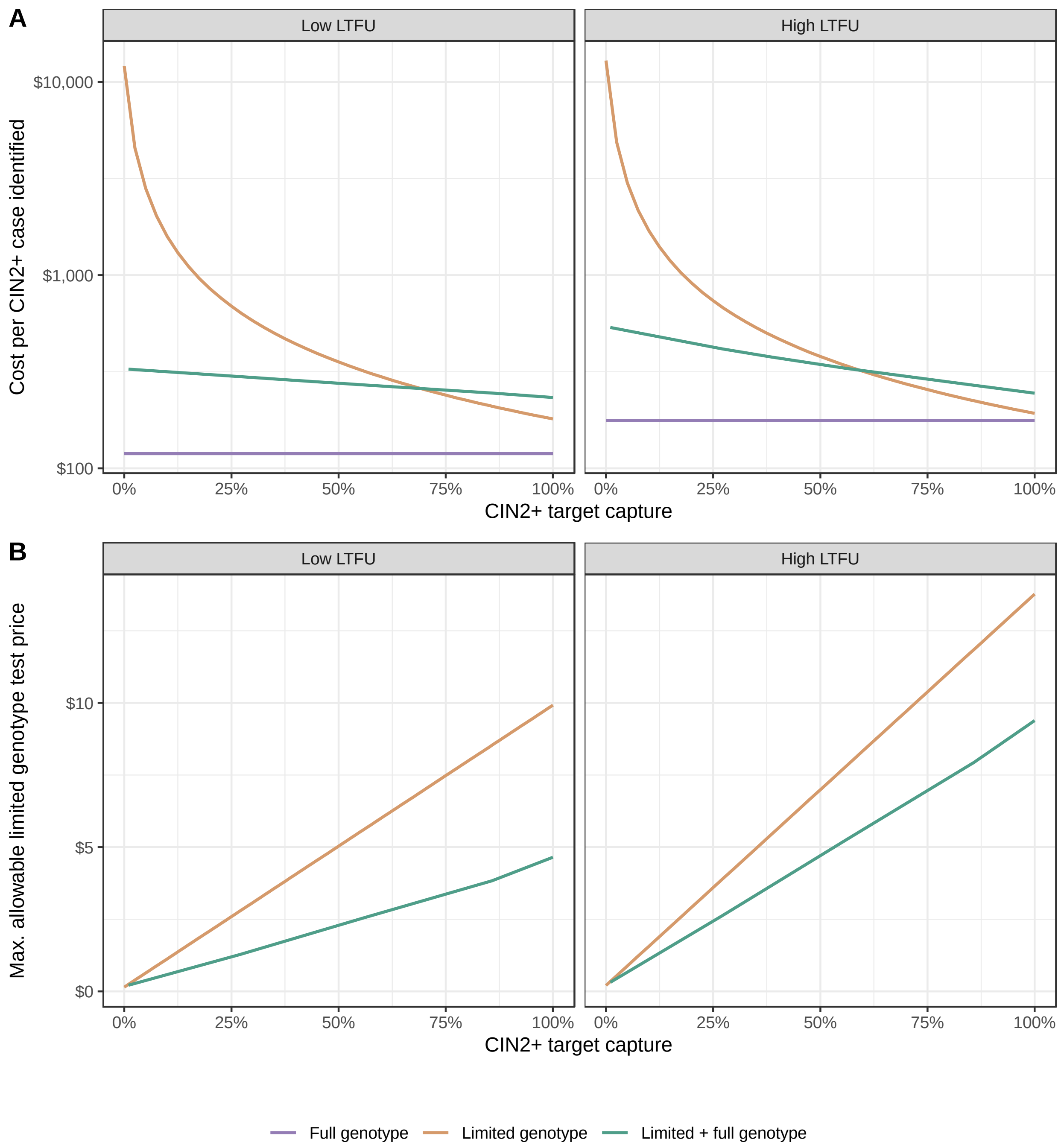


*Figure S5. (A) The cost per CIN2+ case identified (at US $15 per limited-genotype test) and the (B) maximum allowable limited-genotype test price across CIN2+ cases captured in test targets when used alone (Limited-genotype) or with full-genotype testing for those negative by limited-genotype testing (Limited + full genotype), to be cost-equivalent to full-genotype screening (at US $10 per test) under various CIN2+ target capture and loss to follow-up scenarios when disease prevalence is high (55% HPV prevalence, 10% CIN2+ prevalence).*

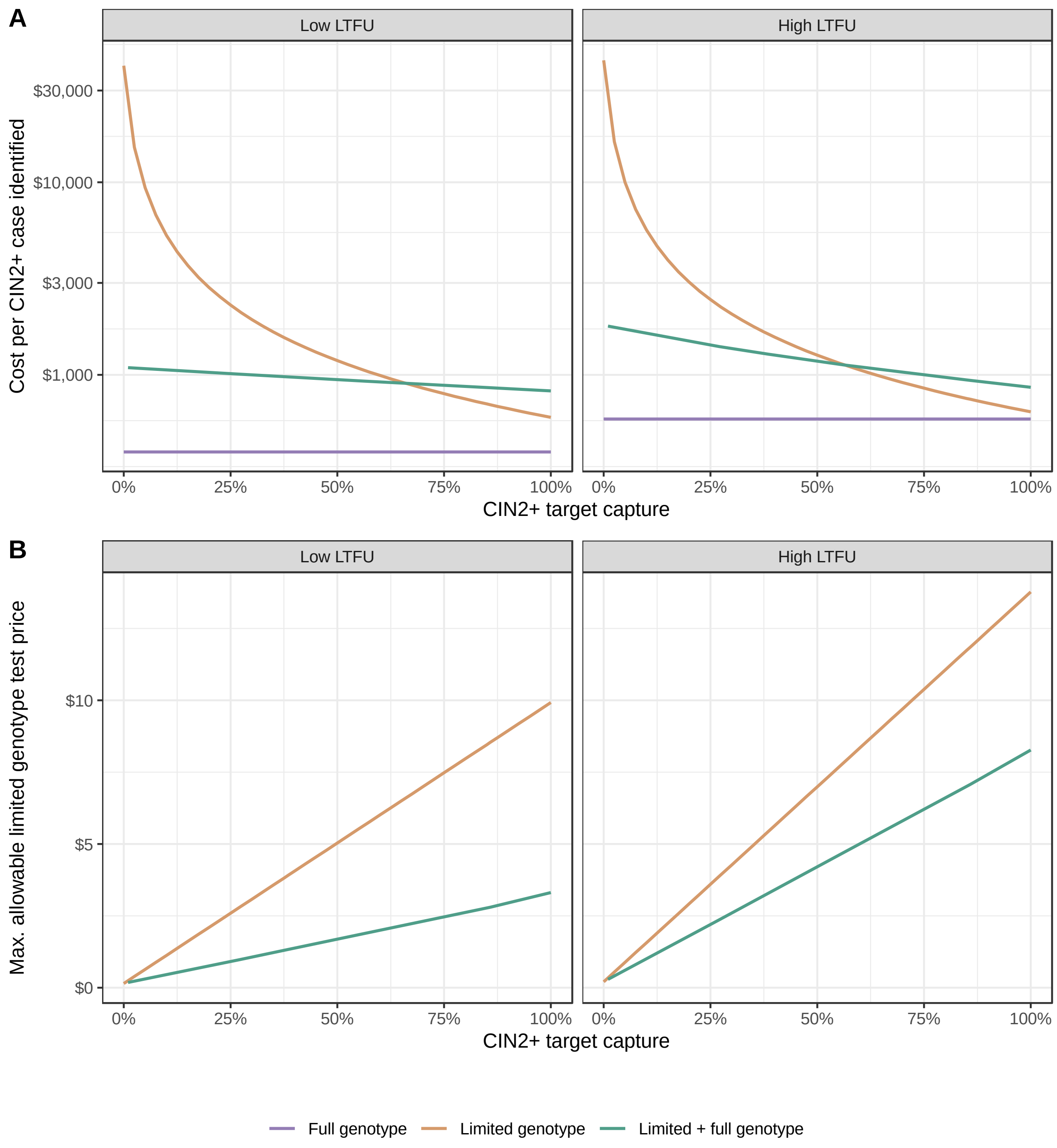


*Figure S6. A) The cost per CIN2+ case identified (at US $15 per limited-genotype test) and the (B) maximum allowable limited-genotype test price across CIN2+ cases captured in test targets when used alone (Limited-genotype) or with full-genotype testing for those negative by limited-genotype testing (Limited + full genotype), to be cost-equivalent to full-genotype screening (at US $10 per test) under various CIN2+ target capture and loss to follow-up scenarios when disease prevalence is high (20% HPV prevalence, 3% CIN2+ prevalence).*

*Table S1. CIN2+ case identification and costs of HPV screening with limited genotype (scenario 1), full-genotype tests (scenario 2), and rule-in limited genotype tests with full genotype testing for those testing negative (scenario 3) when the limited-genotype test costs either US $15 or US $5. The pairwise incremental cost-effectiveness ratio (ICER), estimated for each iteration of scenarios 1 and 3 versus scenario 2, presents the cost of an additional CIN2+ case identified relative to full-genotype testing. Scenarios are denoted as dominated if either less costly and less effective, or more costly and less effective; and denoted as dominant if less costly and more effective*

| | | | | **Limited-genotype test: US $15** | | | **Limited-genotype test: US $5** | | |
|---|---|---|---|---|---|---|---|---|---|
| **Scenario** | HPV targets | CIN2+ target capture | Proportion of CIN2+ cases identified | Total tests costs | Cost per CIN2+ case identified | Pairwise ICER to full-genotype | Total test costs | Cost per CIN2+ case identified | Pairwise ICER to full-genotype |
| **Low loss to follow-up** | | | | | | | | | |
| **Scenario 2 – Full genotype** | **All** | **100%** | 83.9% | US $ 10 000 | **US $ 209** | | US $ 10 000 | **US $ 209** | |
| **Scenario 1 – Limited genotype** | 12 GT | 100% | 83.2% | US $ 15 000 | US $ 316 | dominated | US $ 5 000 | US $ 105 | dominated |
| **Scenario 3 – Limited + full genotype** | 12 GT | 100% | 91.4% | US $ 21 952 | US $ 422 | US $ 2 796 | US $ 11 952 | US $ 230 | US $ 457 |
| **Scenario 1 – Limited genotype** | 8 GT | 86% | 71.6% | US $ 15 000 | US $ 368 | dominated | US $ 5 000 | US $ 123 | dominated |
| **Scenario 3 – Limited + full genotype** | 8 GT | 86% | 88.5% | US $ 22 267 | US $ 441 | US $ 4 634 | US $ 12 267 | US $ 243 | US $ 856 |
| **Scenario 1 – Limited genotype** | 4 GT | 56% | 47.2% | US $ 15 000 | US $ 557 | dominated | US $ 5 000 | US $ 186 | dominated |
| **Scenario 3 – Limited + full genotype** | 4 GT | 56% | 82.5% | US $ 22 642 | US $ 481 | dominated | US $ 12 642 | US $ 269 | dominated |
| **Scenario 1 – Limited genotype** | 2 GT | 39% | 32.9% | US $ 15 000 | US $ 800 | dominated | US $ 5 000 | US $ 267 | dominated |
| **Scenario 3 – Limited + full genotype** | 2 GT | 39% | 79.0% | US $ 22 875 | US $ 508 | dominated | US $ 12 875 | US $ 286 | dominated |
| **High loss to follow-up** | | | | | | | | | |
| **Scenario 2 – Full genotype** | **All** | **100%** | 56.6% | US $ 10 000 | **US $ 310** | | US $ 10 000 | **US $ 310** | |
| **Scenario 1 – Limited genotype** | 12 GT | 100% | 77.9% | US $ 15 000 | US $ 338 | US $ 411 | US $ 5 000 | US $ 113 | dominant |
| **Scenario 3 – Limited + full genotype** | 12 GT | 100% | 82.5% | US $ 20 785 | US $ 442 | US $ 730 | US $ 10 785 | US $ 229 | US $ 53 |
| **Scenario 1 – Limited genotype** | 8 GT | 86% | 67.0% | US $ 15 000 | US $ 393 | US $ 840 | US $ 5 000 | US $ 131 | dominant |
| **Scenario 3 – Limited + full genotype** | 8 GT | 86% | 76.5% | US $ 21 048 | US $ 483 | US $ 972 | US $ 11 048 | US $ 253 | US $ 92 |
| **Scenario 1 – Limited genotype** | 4 GT | 56% | 44.2% | US $ 15 000 | US $ 595 | dominated | US $ 5 000 | US $ 198 | dominated |
| **Scenario 3 – Limited + full genotype** | 4 GT | 56% | 64.0% | US $ 21 359 | US $ 585 | US $ 2 668 | US $ 11 359 | US $ 311 | US $ 319 |
| **Scenario 1 – Limited genotype** | 2 GT | 39% | 30.8% | US $ 15 000 | US $ 855 | dominated | US $ 5 000 | US $ 285 | dominated |
| **Scenario 3 – Limited + full genotype** | 2 GT | 39% | 56.7% | US $ 21 553 | US $ 667 | US $ 175 641 | US $ 11 553 | US $ 358 | US $ 23 610 |
